# Disability inclusion in the Brazilian health system: results of a health system assessment

**DOI:** 10.1101/2024.06.04.24308469

**Authors:** Sarah Polack, Vinicius Delgado Ramos, Luciana Sepúlveda Köptcke, Indyara de Araujo Morais, Veronika Reichenberger, Nathaniel Scherer, Maria do Socorro Veloso de Albuquerque, Hannah Kuper, Tereza Maciel Lyra, Christina May Moran de Brito

**Author notes:** Corresponding author: Sarah Polack, International Centre for Evidence in Disability, London School of Hygiene & Tropical Medicine, Keppel Street, London WC1E 7HT, United Kingdom.

## Abstract

**Background:** People with disabilities face more barriers accessing healthcare and, on average, experience worse health outcomes. Strengthening health access for people with disabilities requires coordinated action across the health system. The Missing Billion Inclusive Health System Framework is a new tool to support policy makers assess levels of disability inclusion within health systems. In this study we use the framework within the Unified Health System in Brazil. We consider the relevance and feasibility of the indicators, as part of further testing and refining the framework.

**Methods:** Information sources, used to complete the assessment, included Brazilian laws and policies, publically available data, published literature and interviews with people with disabilities and service providers. A workshop with stakeholders was held to co-develop key recommendations.

**Findings:** Overall, the framework was comprehensive and feasible to complete. It highlighted key strengths in terms of disability inclusion in the Brazilian health system as well as gaps and leverage points for action.

**Interpretation:** The Missing Billions framework can identify progress and opportunities to strengthen disability inclusion in health systems. In Brazil, key promotive factors include supportive policies, leadership and financing structures. There are also opportunities for strengthening data and evidence, healthcare worker training on disability and health service accessibility. Actions must be centered on, and informed by, people with disabilities.

**Funding:** This work was supported by the São Paulo Research Foundation, Brazilian National Council for Scientific Technological Development, Federate District Research Foundation and the Medical Research Council. Hannah Kuper is supported by a NIHR Global Research Professorship.

## INTRODUCTION

Health is a disability rights issue; Article 25 of the United Nations Convention on the rights of Persons with Disabilities, to which Brazil is as a signatory, states that “persons with disabilities have the right to the enjoyment of the highest attainable standard of health without discrimination on the basis of disability”.^1^ Access to healthcare for people with disabilities is also a development issue as an estimated 16% of the population live with a disability,^2^ this is crucial for achieving Universal Health Coverage and the Sustainable Development Goals (SDGs) on health, and targets that are dependent on heath. However, evidence suggests we are falling short of achieving inclusive healthcare. People with disabilities are more likely to face barriers in accessing quality and appropriate health care^3^, despite having, on average, higher health care needs, related to their underlying impairment and health condition.^3,4^ Barriers may include lack of accessible information or sign-language interpreters, physically inaccessible buildings or transport to health services, and negative attitudes, and stigma by health care professionals^4^. These barriers often result from systems failures such as a lack of disability inclusive policies. People with disabilities are also at greater risk of economic exclusion and poverty,^5^ often compounded by ‘extra-costs of disability’ (e.g. need for assistive technology, accessible transport);^6^ therefore, financial barriers to health-care are also common. The consequence of higher health needs, coupled with widespread barriers to access, are evident in poor health outcomes data. For example, according to literature reviews and analysis of national datasets, people with disabilities have two-fold higher mortality rate, are two-times more likely to have diabetes and are five-times more likely to experience catastrophic health expenditure compared to people without disabilities.^3,4,7,8^

Strengthening health access for people with disabilities is therefore important and will require coordinated action across all levels of the health-system. The focus is often put on improving service level components (e.g. physical accessibility of services, health staff knowledge, and attitudes). However, these issues are typically driven by deeper system failures, such as weak policies, insufficient commitment to disability inclusion within leadership, lack of knowledge, and insufficient financing. Therefore, a whole system level approach is needed. As the situation will vary between countries, there is a need, at national levels, to understand what is happening, what is working well and what the gaps and opportunities are in relation to disability inclusion across the health system. The Missing Billion Inclusive Health System Framework^7^ (Figure 1), co-developed by the Missing Billion Initiative and key global stakeholders, may be useful for the assessment of disability-inclusion in a national health system. The framework describes nine key health system components: governance, leadership, health financing, data and evidence, autonomy and awareness, affordability, human resources, health facilities, and rehabilitation services and assistive technology (AT). For each component, there is a set of indicators (see Tables 1–3 and Supplementary Material Tables 1-3) to identify progress and gaps and, ultimately, inform and monitor action towards better disability inclusion. It is intended to be conducted by a task team that includes relevant stakeholders from the Ministry of Health, people with disabilities and their representative organisations, as well as nongovernmental organisations (NGO) and technical partners.

**Figure 1.**
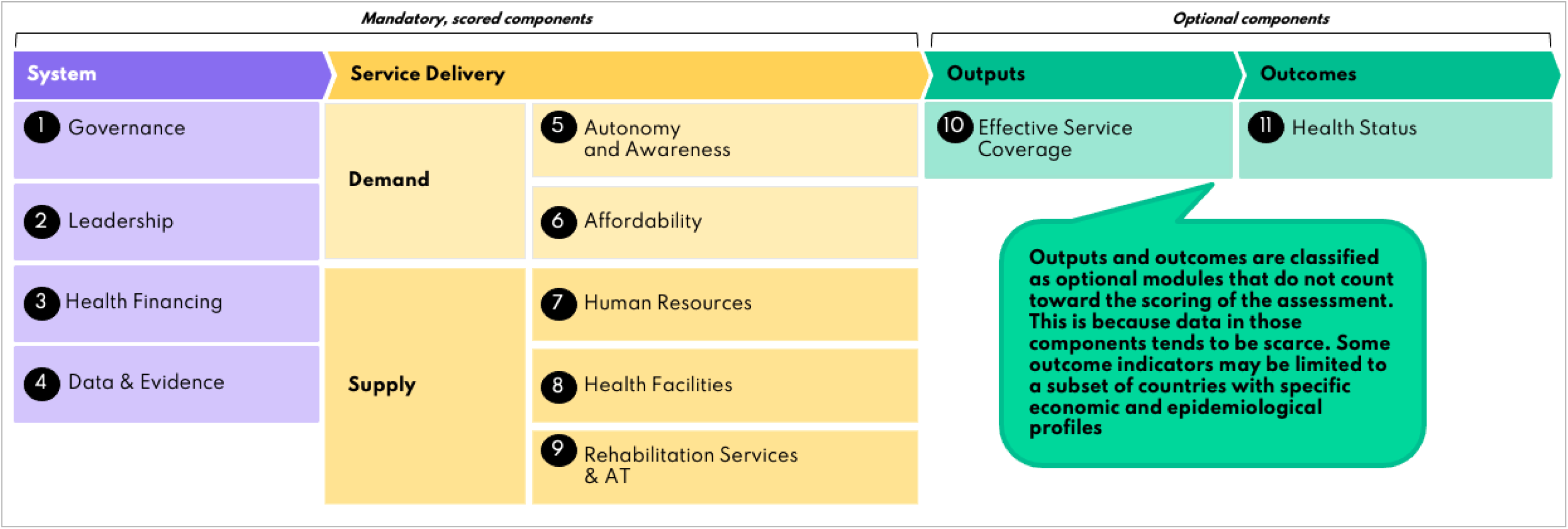
Missing Billion Disability-Inclusive Health System Framework ^7^

**Table 1:** Systems levels indicators, status and score.

| Indicator and definition | Status in Brazil | Score |
| --- | --- | --- |
| **1. GOVERNANCE** |  |  |
| 1.1 Ratification of UNCRPD | **Yes** UNCRPD ratified in 2018 and evidence of it being actioned | **1** |
| 1.2 Existence of national law protecting rights of persons with disabilities to health | **Yes** 2023 National Plan for the Rights of Persons with Disabilities, Decree No 11.793 and Brazilian Inclusion Law/Law 13,146. These laws<br>1) prohibit discrimination in healthcare and<br>2 )require reasonable accommodation for people with disabilities | **1** |
| 1.3 Existence of national policy or decree on health for persons with disabilities | **Yes.** 2023 National Policy for Comprehensive Health Care for People with Disabilities. (see **Appendix 1**). Policy ensures:<br>1) general healthcare services for persons with disabilities<br>2) access to rehabilitation, other specialists and assistive technology services,<br>3) measures to implement these. | **1** |
| 1.4. Inclusion of people with disabilities in National Health Sector Plan(s) | **Partially.** National Health Plan 2020-23 includes:<br>1) Some actions and targets for persons with disabilities within dental care services<br>2) Actions and targets for specialist health services for persons with disabilities:<br>It does not include:<br>3) Basic statistics about persons with disabilities and health<br>4) Monitoring and evaluation indicators on disability as part of overall framework for the health sector | **0.5** |
| 1.5 Inclusion of people with disabilities in National disease plan (e.g., HIV, rare diseases, hepatitis) | **Partially.** Not mentioned in the HIV plan. The Rare Diseases Policy Guidelines provides some guidance on how primary and secondary healthcare services should refer people with rare diseases and disabilities. | **0.5** |
| 1.6 Cross-ministry taskforce or structure to coordinate work on disability inclusion | **Yes.** There is a "Cross-Sector Commission on the Healthcare of Persons with Disabilities" which includes the Ministry of Health | **1** |
| **COMPONENT SCORE** |  | **0.8 (HIGH)** |

| **2. LEADERSHIP** |  |  |
| --- | --- | --- |
| 2.1 MoH Leadership: Existence of a focal point/team in MoH that's responsible for ensuring health access for people with disabilities | **Yes:** General Coordination for the Health of People with Disabilities is responsible for implementing national health policy for people with disabilities (including general healthcare and rehabilitation). | **1** |
| 2.2 National health sector coordination: Formal representation of persons with disabilities in highest-level health sector **coordination** structures | **Yes:** OPDs are represented among "user groups" within the National Health Council; National Disability Rights Council considers health-related issues | **1** |
| 2.3 Pandemic preparedness structure: Formal representation of people with disabilities (individuals or OPD) in national taskforce (e.g. COVID) | No evidence that people with disabilities or OPDs are mandatorily or systematically included in the two main structures to coordinate preparedness and response to public health emergencies (Events Monitoring Committee and the Health Emergencies Operations Center). No evidence that persons with disabilities were represented within the three main COVID coordination groups | **0** |
| **COMPONENT SCORE** |  | **0.7 (MEDIUM)** |
| **3. HEALTH FINANCING** |  |  |
| 3.1 Disability Inclusion Budget: Budget (MoH or devolved level) for role/department in MoH working on disability inclusion | **Yes.** All three levels of government (municipal, state and federal administration) share the responsibility to fund the public healthcare system. They pay for their own initiatives and also transfer funds between them. | **1** |
| 3.2 Reimbursement adjustment for services provided to patients with disabilities | **Yes.** Brazil has universal public healthcare system (Unified Health System) which offers health and rehabilitation services free at the point of care.<br>For people who use private healthcare Laws 9656/1998 and 14454/2022 establish that: private healthcare insurance cannot discriminate clients on the basis of disability (art 14) and must cover any procedure that has its effectiveness scientifically proved, is recommended by the SUS National Commission on the Adoption of Technology, or recommended by at least on internationally renowned health technology assessment body. | **1** |
| 3.2 Funding for AT/rehabilitation in MoH (or devolved levels) budget | **Yes.** Estimates, using 2019 MoH data, suggest ~1 % of total government health expenditure was related to rehabilitation (at secondary and tertiary level) and 0.36% to APs. | **1** |
| **COMPONENT SCORE** |  | **1 (HIGH)** |

| **4. DATA AND EVIDENCE** |  |  |
| --- | --- | --- |
| 4.1 Maturity of disability and health data collection | 2022 Census and 2019 National Health Survey included questions on disability (adapted from the Washington Group questions) which would allow disability disaggregation of health data. Electronic health information records can be disaggregated by 2 or 3 digits ICD codes relating to health conditions and some impairments, and only indirectly to disability. No detailed disability identification method available. | 0.33 |
| 4.2 Quality of disability and health data collection method | For census/Surveys:<br>1) Data collection method is valid<br>2) Data collection is recent (2019 and 2022)<br>3) Data collection is nationally representative<br>4) More than 5 impairment (functional domains) types are covered | 1 |
| 4.3 Maturity of disability and health data usage | Unknown; Disability-disaggregated socio-economic indicators (education, income, occupation) reported, but no evidence found of analysis of disability and health data from the NHS/census | 0 |
| 4.4 Quality of disability and health data usage method | Unknown; (see 4.3) | 0 |
| **COMPONENT SCORE** |  | **0.3 (LOW)** |

The Missing Billion framework has been pilot-tested in Zimbabwe and the Maldives and indicators were revised based on lessons learnt.^9^ The updated version requires further testing in different countries. Brazil provides an important opportunity for this type of assessment. It is the largest country in South America and has progressive laws demonstrating commitment to disability inclusion, including within health. Brazil has a universal public healthcare system, called Sistema Único de Saúde (SUS) and a National Health Policy for People with Disabilities initiated in 2002, and updated in 2023, specifically to address health inequalities. However, a recent scoping review highlighted that despite supportive policies, there are substantial implementation gaps and therefore inequalities in healthcare access persist for people with disabilities.^10^

This study aimed to apply the Missing Billion’s framework in Brazil in order to: i) assess the acceptability, feasibility and usefulness of the framework indicators in this setting; ii) understand the extent of disability inclusion in the health system and key gaps; and iii) inform the co-development of key recommendations for action with key stakeholders.

## METHODS

This work was conducted as part of a four-year research project exploring access to health services for people with disabilities in Brazil. This project was conducted within a research partnership between the University of São Paulo, Fundação Oswaldo Cruz (Fiocruz) and the London School of Hygiene & Tropical Medicine.

The Missing Billion Inclusive Health Systems Framework (see Figure 1) was used to structure a situational analysis^7^. Each component within ‘Health System’ and ‘Service Delivery’ has set of indicators and each indicator has a definition, information required and scoring metric (See Tables 1–2 and Supplementary material Tables 1-2)^11^. An average score is calculated for each component. Scores of >0.75 are defined as ‘high’, 0.5-0.74 as ‘medium’ and <0.5 as ‘low’. The ‘outputs’ and ‘outcomes’ components are optional modules and do not have scoring metrics as these data tend to be scarce.

**Table 2:**
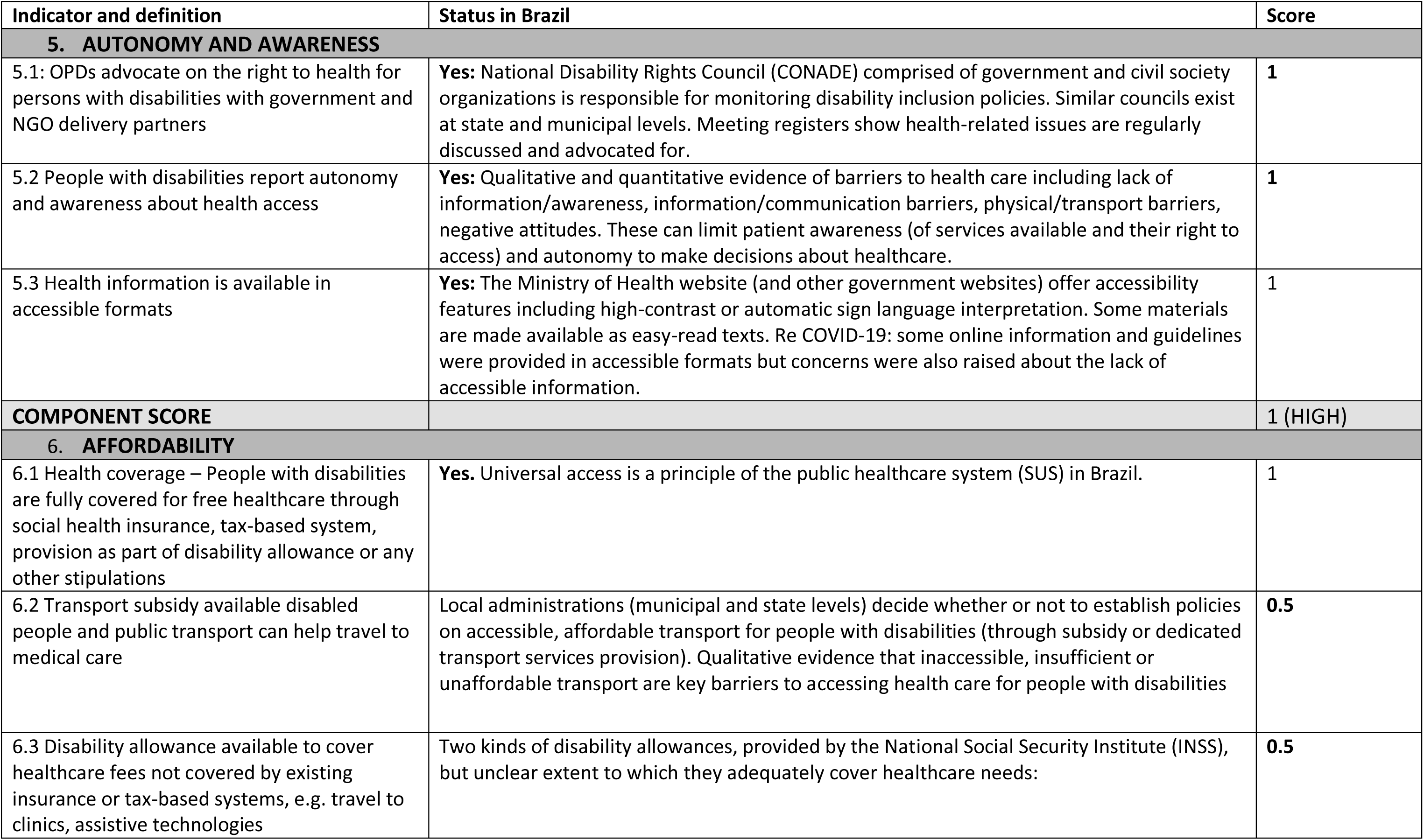
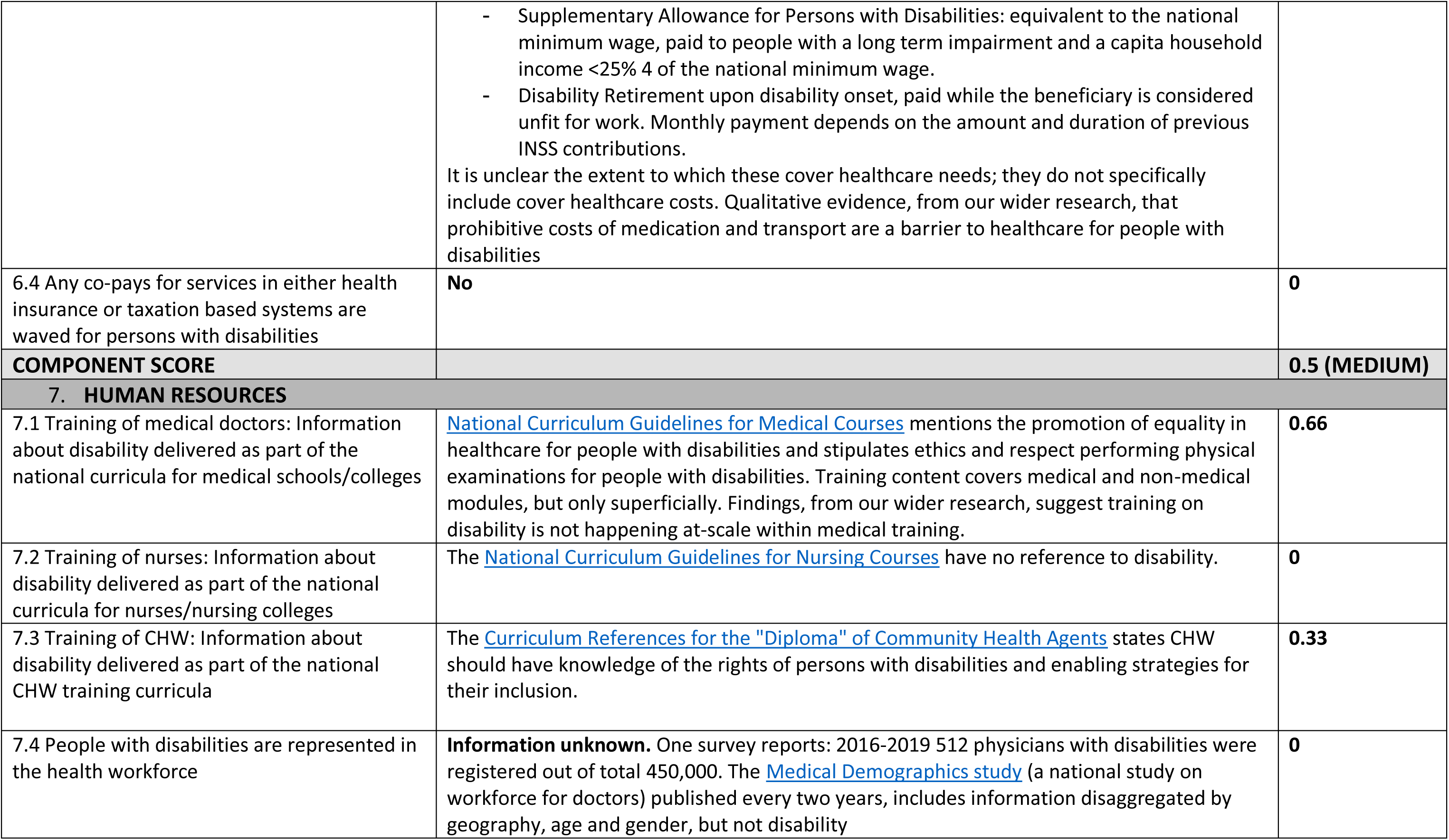
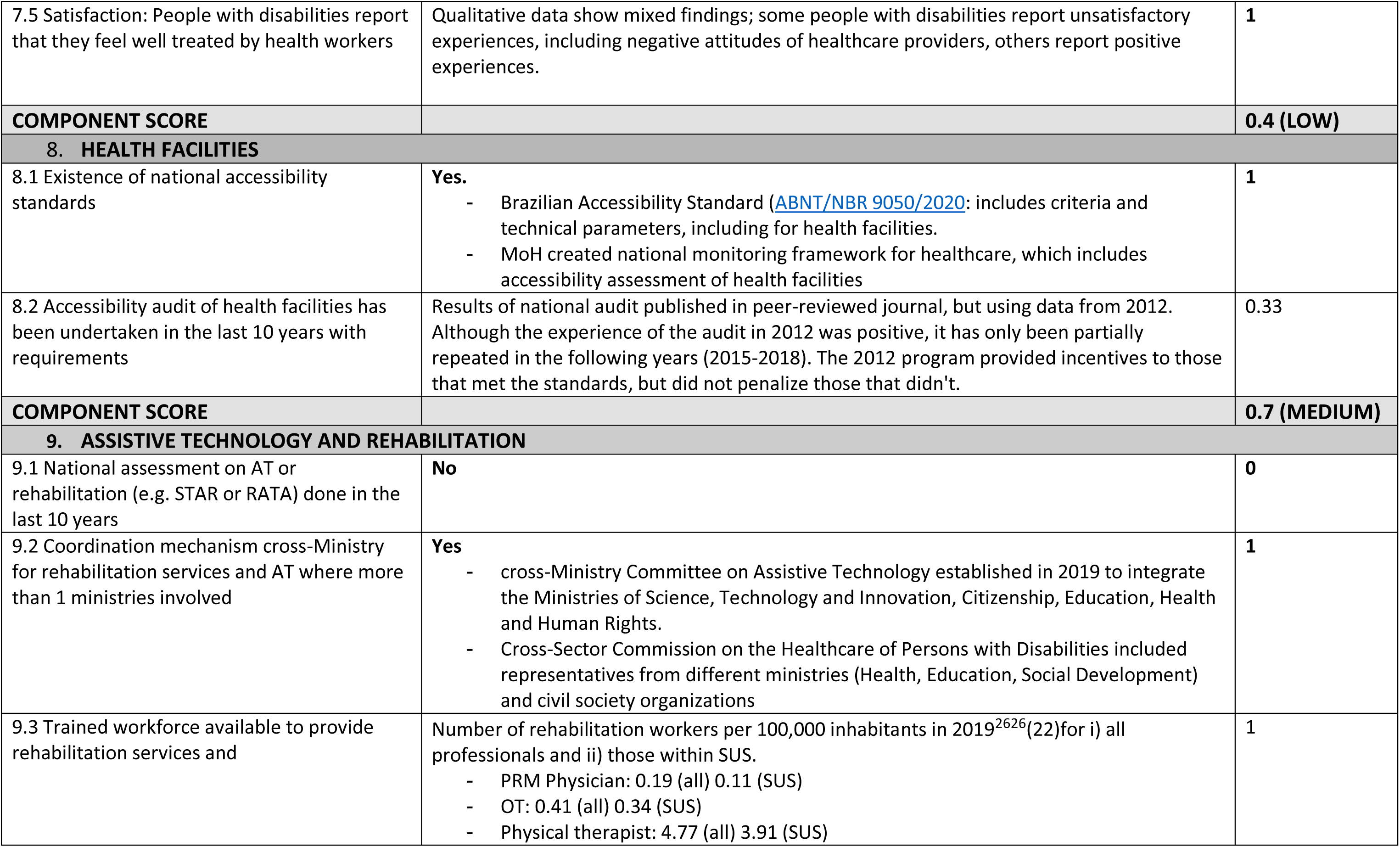
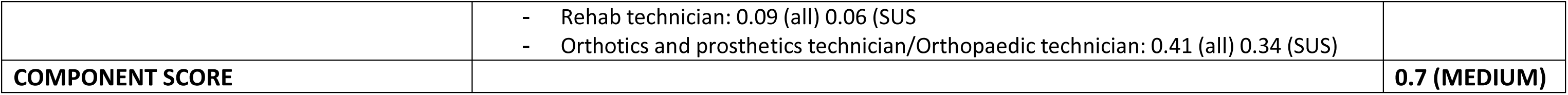
Service delivery levels indicators, status and score.

The framework is based on the WHO Building Blocks and the Primary Health Care Performance Initiative framework^12^. The framework and indicators were reviewed by a range of experts (governmental and UN stakeholders, health systems specialists, academics and disability rights organizations). They were pilot tested in the Maldives and Zimbabwe and the framework was subsequently refined.^9^

The intention, for this framework, is that the assessment is led by the Ministry of Health of the respective country. However, for this study, it was conducted, by a team of academics from Brazil and UK, including Brazilian health and rehabilitation practitioners, to further pilot test and refine the tool. The assessment was completed between November 2022-March 2023 and updated in November-Feb 2023.

We used several sources to collect information for the indicators including: i) Brazilian policies and laws (see supplementary material table 4); ii) publicly available data (e.g. from Brazilian health information systems); iii) published literature, including peer reviewed journal articles; and iv) information known to the Brazilian authors, including public health and rehabilitation professionals and academics with expertise in disability inclusion. We also drew on the findings from our wider study on access to health services for people with disabilities which has included: in-depth interviews with 87 people with disabilities and 57 service providers from São Paulo, Santos, Brasília and Arcoverde (reported in a separate forthcoming publication), an analysis of a national health services accessibility audit,^13^ a scoping review of the evidence of disability inclusion in the health system in Brazil,^10^ an analysis of and a rehabilitation system situational analysis (reported in separate forthcoming publication).

For each indicator, co-authors also reflected on clarity, relevance and the feasibility of collecting the required information, as part of further testing and refining of the framework.

### Co-development of recommendations

A final study meeting was held in Brasilia, Brazil in March 2023 to share findings from the research, including the health system assessment. This included a participatory workshop to co-develop recommendations on disability inclusion informed by the study findings. Participants included researchers, representatives from organisations of people with disabilities, and national and subnational executive, legislative and judicial branches of government, health and rehabilitation service providers. Key findings from the research were shared and discussed with workshop participants. Guiding questions were posed to workshop participants to stimulate discussion about implications of the findings and, through this process, six key recommendations were agreed upon.

#### Ethical approval

Ethical approval was obtained for this research from the ethics committees of the London School of Hygiene & Tropical Medicine, the University of São Paulo Medical School General Hospital and the São Paulo Municipal Health Department, and Fiocruz branches in Brasilia and Recife.

## RESULTS

Details on the indicator metrics and scoring system for the Missing Billion framework are provided in Supplementary tables 1-3. We summarise the findings for each indicator at health system (Table 1), service level (Table 2) and outputs/outcomes (Table 3), and reflect on the use of the indicators themselves. Next, we present key recommendations arising from the participatory workshop.

**Table 3.** Health outputs and outcomes indicators and status in Brazil.

| Indicator and definition | Status in Brazil |
| --- | --- |
| **9. EFFECTIVE SERVICE COVERAGE** |  |
| 10.1 Modern contraception coverage: Women whose demand is satisfied with modern method of contraception disaggregated by disability | **Information unknown.** 2019 NHS collected data on disability and types of contraception used; but doesn't ask whether demand is satisfied |
| 10.2 ART coverage: People with HIV receiving ART, disaggregated by disability | **Information unknown** |
| 10.3: DPT coverage: Children aged 12-23 months who have received diphtheria-tetanus-pertussis vaccine (DTP3), disaggregated by disability | **Information unknown** |
| 10.4 Refractive error coverage: People with refractive error have coverage of glasses | **Information at national level unknown.**<br>- Eye survey in Brazilian Amazon Region in 2014-15 found 29% and 30% effective coverage for distance and near refractive error respectively |
| 10.5 NCD coverage: People with diabetes on treatment OR people with hypertension on treatment, disaggregated by disability | **Information at national level unknown.**<br>- Survey in Padre Paraíso, Minas Gerais state and Poções, Bahia state: Among people with known diabetes and hypertension more than 93% of people with and without physical disabilities reported access to treatment (article submitted ) |
| **10. HEALTH STATUS** |  |
| 11.1: Overall mortality rate, disaggregated by disability | According to the Missing Billions data dashboard, the Mortality Rate is<br>- 1.3-1.7 times higher for people with disabilities than people without disabilities (based on general/functional definition of disability)<br>- 1.7 times higher for people with mobility impairment than for people without<br>• 1.74 times higher for people with depression<br>• 1.7 times higher than for people with cognitive impairment |
| 11.2 Prevalence of diabetes OR hypertension among persons aged 18+ years, disaggregated by disability | **Information unknown**<br>- National: 2019 NHS collects data on diabetes, hypertension and disability, so potential to calculate |
| 11.4: Prevalence of HIV, disaggregated by disability | **Information unknown** |
| 11.5 Prevalence of overweight and obesity among persons aged 18+ years, disaggregated by disability | **Information unknown at national level**<br>- Survey in Padre Paraíso, Minas Gerais state and Poções, Bahia state found: Overweight: 36% of adults with and without physical disability; Obesity: 11% adults with physical disability; 16% to adults without physical disabilities |
| 11.6: Wasting: prevalence of children wasted) disaggregated by disability | **Information unknown** |

### Systems level indicators (Table 1)

#### Governance

Indicators 1.1-1.4 highlight that Brazil has a progressive policy framework committed to disability inclusion. For more information, see Supplementary Table 4 which summarises key policies and content related to disability and health. The UNCRPD was ratified in 2008 and national disability-focussed laws/decrees prohibit discrimination on the basis of disability and promote the rights of persons with disabilities to health. In 2023, a revised National Health Policy for Persons with Disabilities (originally produced in 2002) was published called the National Policy on Comprehensive Health Care for Persons with Disability (PNAISPD). This includes implantation guidance on disability inclusion in general health care and specialist services as well as specifications regarding budget, monitoring and responsibilities of actors.

Some governance gaps were highlighted. The recent National Health Plan (2020-23) includes some targets for people with disabilities.^14^ However, these typically focus on specialist services (e.g., related to vision and mobility), rather than inclusion in general health services and monitoring and evaluation indicators related to disability appear lacking. The Rare Diseases Policy Guidelines provides some guidance on how primary and secondary healthcare services should refer people with rare diseases and disabilities. However, there is no explicit consideration of people with disabilities in the National HIV plan.

The framework indicators for governance were considered relevant and straightforward to answer; most of the information was available in the public domain, through policy documents, or known to the Brazilian authors. We recommend graded scoring, rather than binary, for indicator 1.5 (Inclusion of people with disabilities in National disease plan) as it may vary, as in the case of Brazil where disability was mentioned in some plans (e.g. rare diseases) and not others (e.g. HIV)

#### Leadership

The General Coordination for the Health of Persons with Disabilities, within the Ministry of Health, is responsible for implementing disability-related health policies. Further, Organisations of Persons with Disabilities (OPDs) are represented within the Participatory Health Councils. These are Collegiate bodies, with both deliberative and consultative roles, established at municipal, state and federal levels, in which representatives of the public administration and civil society participate to implement and monitor public health policies.

However, the General Coordination for the Health of Persons with Disabilities appears to have a primary focus on specialist services (e.g. rehabilitation, including assistive technology) and priorities relating to general health services are less clear. Indicators on pandemic preparedness, highlighted leadership-related gaps; there is no evidence of formal representation of people with disabilities or OPDs in current national taskforces or the previous COVID-19 specific committees/taskforces.

The three leadership indicators for governance were considered relevant and straightforward to answer.

#### Financing

The Unified Health System in Brazil offers health and rehabilitation services free at the point of care. There is financial commitment to supporting the health of persons with disabilities, through the General Coordination for the Health of Persons with Disabilities. Specifically, federal funds are available to invest in infrastructure and expansion of rehabilitation facilities, neonatal screening and adapted vehicles to be used by health services (see Supplementary Table 4). Although a scoping review, highlighted challenges with this, including insufficient funding and delays in implementation.^15^ Estimates from 2019 suggest that approximately 1% of government funding was allocated to rehabilitation at secondary and tertiary level and 0.36% to assistive products (AP) provision^16^. Considering global estimates, that a third of people are expected to need rehabilitation services, this seems low ^17^ and research has highlighted issues of chronic underfunding of rehabilitation in Brazil.^15^

The financing indicators were generally clear. The guidance for indicator 3.2 (on funding for AT/rehabilitation) could be clarified to indicate how information on budget allocation should be taken into account in the scoring.

#### Data and Evidence

Findings relating to data and evidence indicators are mixed. Questions on disability were included in the latest National Health Survey (NHS; 2019)^18^ and Census (2022) which would allow disaggregation of collected health data by disability status. We were not able to find evidence of such analysis having been conducted. Brazil has a national health data and information systems; with electronic health records being progressively implemented for the whole population. These administrative and clinical datasets have vast potential for understanding health needs of the population. However, to date, no disability markers are included in the dataset, prohibiting disability data disaggregation.^10^

The data and evidence indicators were considered generally clear, relevant, and appropriate. It may be helpful to clarify that ‘disability and health data’ refers to data on disability and access to general health services and health outcomes, not just specialist services.

### Service level indicators (Table 3)

#### Autonomy and awareness (demand)

Organisations of Persons with Disabilities (OPDs) discuss and advocate on issues of health access, in particular the National Disability Rights Council (a participatory council comprised of government and civil society organisations but dedicated to disability policies).^19^

We did not find direct data on autonomy in the context of healthcare. However quantitative and qualitative studies^10^ find evidence of unmet healthcare needs and barriers that limit the autonomy of people with disabilities to make decisions about their healthcare (e.g. inaccessible information, communication and physical barriers). In terms of information accessibility, the Ministry of Health website offers some accessibility features including high-contrast or automatic sign language interpretation and easy read text for some materials. Specifically for COVID-19, while some online information and guidelines were provided in accessible formats, concerns were also raised about the lack of accessible information^20,21^. Qualitative data, collected as part of our wider study, highlighted information barriers particularly for people with visual impairment; for example, medication information that is not available in Braille prevents checking of expiry dates.

Indicators related to autonomy and awareness were generally considered appropriate, and the eligibility of both quantitative and qualitative information sources was appreciated. Autonomy is a multi-dimensional concept and data directly assessing autonomy to make informed choices about health care may not be widely available, although measurement of this deserves attention in future research. Therefore, within the framework (indicator 5.2), it may be helpful to encourage inclusion of information on likely (personal/contextual) factors that influence autonomy (e.g., transport, costs, information/communication, cost, as well as awareness of rights). Further, the phrase ‘Awareness about health access’ is somewhat vague and potentially confusing; more specific examples may be helpful, e.g. awareness of their rights, awareness of services that are available. There is also a discrepancy in the requirements for quantitative (data collected) and qualitative (results published) sources; we recommend that to score 1, data from either source should be analysed and published.

#### Affordability (demand)

Universal access is a principal of the public healthcare system in Brazil, with health-care free at the point of use. However, qualitative evidence in Brazil suggests, including from our wider research, that out of pocket payments for transport, AT, and medications can be a major barrier for people with disabilities, who are likely to have higher needs for health care and are, on average, poorer.^5,6,22^ Further, limitations with public services mean some people seek private care thus incurring extra costs, which may be particularly high for people with disabilities considering greater healthcare needs.

There are two disability allowances in Brazil provided by the National Social Security Institute (INSS). The Supplementary Allowance for Persons with Disabilities (Benefício de Prestação Continuada, BPC) pays the equivalent to the national minimum wage to people with long-term impairments and a household per capita income less than 25% of the national minimum wage. The coverage and extent to which these allowances, cover additional health costs (e.g. AT, accessible travel) incurred by people with disabilities is unknown.

Affordability indicators were generally feasible to address. All the indicators in this domain refer to state provisions, allowances and health funding systems. However, they don’t capture affordability from the perspective (experience) of people with disabilities. Cost (direct and indirect) is widely reported as a barrier in both quantitative and qualitative research on health access. Therefore, it may be helpful to include an additional indicator, similar to 5.2, on published evidence about affordability from qualitative or quantitative data.

#### Human resources (supply)

There is no mention of disability in the National nurse training guidelines and limited mention in the National Curriculum guidelines for Medical doctors and Community Health workers. This focus is limited to statements about the importance of awareness on disability rights and inclusion and equality in healthcare for people with disabilities.

Data on disability representation in the health workforce was also lacking. There is the potential to generate this information considering data on the doctor workforce, by age and gender, are published every two years. We were unable to find quantitative data comparing satisfaction with healthcare workers between people with and without disabilities. However, there is some qualitative evidence of attitudinal barriers to health care; with people with disabilities reporting negative attitudes from healthcare staff.^10,23^ We recommend that scoring for indicator 7.3 (representation of people with disability in the health workforce) includes an option of ‘0’ for where this information is not collected/available. The score for indicator 7.5 could be misleading (‘Satisfaction: whether people with disabilities report they feel treated well by health workers’) as a score of 1 is assigned if data are present, even if the data suggest people are treated poorly. Similarly, a medium-score was allocated for training of medical doctors, based on criteria, but our wider research indicates that the actual content on disability-inclusive healthcare is very superficial.

#### Health facilities (supply)

Brazil has Accessibility Standards for public buildings, including for health facilities. There is also a national monitoring framework that includes healthcare accessibility assessment. This was implemented as part of a pay for performance programme (PMAQ) which has now been disbanded. A national audit was conducted in 2012 by trained staff at all 38,812 primary healthcare facilities.^13^ This large audit found that overall accessibility, of internal and external spaces, was generally low and revealed socioeconomic inequalities, with accessibility being generally better in larger (and therefore likely urban) municipalities. The audit highlighted particular accessibility gaps for people with vision and hearing impairments.^13^

It would be helpful to clarify whether the accessibility audit indicator (8.2) refers only to national level or also sub-national levels and how to assign scores accordingly. For example, what score should be assigned if there is evidence of an audit in only one or two health facilities?

#### Rehabilitation services / AT (Supply)

Data on use of rehabilitation services is collected in the National Health Survey; 16% of people with some functioning difficulty reported using rehabilitation services and a secondary analysis of the 2013 NHS data identified social inequities in use of rehabilitation services.^24^ A rapid Assessment of Assistive Technology conducted among people attending rehabilitation services in São Paulo Brazil found high AT need.^25^ However, we did not find evidence of national AT assessments and data on population level rehabilitation and AT need are generally lacking.

In terms of leadership, there is Cross-Ministry Committee on AT; a coordination mechanism for the different ministries involved in AT as well as Cross-sector Commission on health care (including rehabilitation) of persons with disabilities. Data on the rehabilitation/AT workforce indicate that, physical therapists are most commonly available (3.91 within SUS per 100,000 population), followed by Occupational Therapists (0.34 per 100,000 population).^26^

The indicators were generally considered useful and straightforward. For indicator 9.3 it needs to be clearer whether the information required for refers to physiotherapists only or also includes the different workforce listed.

### Health outputs and outcomes (Table 4)

We were unable to identify national level data or estimates on health service coverage (contraception, ART, DPT, diabetes and hypertension treatment) disaggregated by disability. Some sub-national data exist.^27^ The Missing Billion data dashboard indicates higher mortality rate among people with disabilities.^28^ However, data on other health status indicators were lacking. Some these data could be generated using the latest NHS, but we were unable to find evidence of these analysis.

The indicators were clear, though data were generally lacking to answer them. Adding indicators on coverage of AT (besides glasses) and rehabilitation should be considered.

### Co-developed recommendations

Based on these findings, the following six recommendations for improved disability inclusion in the healthcare system were co-developed at the final study workshop:

1. Strengthen healthcare worker training on disability
2. Improve data on disability with respect to health in Brazil
3. Strengthen accessibility of healthcare facilities
4. Increasing availability of, and pathways to, rehabilitation and specialist services
5. Build movement of people with disabilities on access to healthcare; greater advocacy by people with disabilities)
6. Strengthen inter-sectoral policies; recognising barriers to health exist outside of the health system (e.g. transport or urban plan. Not health service alone]

## DISCUSSION

This paper assessed disability inclusion across components of the health system in Brazil. Examples of good practise, that other countries could learn from, include the presence of a General Coordination for the Health of People with Disabilities within the Ministry of Health and the progressive rights-based policies, including the National Policy on Comprehensive Health Care for Persons with Disability and the Brazilian Law for Inclusion. Another important initiative, lacking in other settings, was the national accessibility audit of primary health facilities conducted in 2012.^13^

Our assessment also highlighted some areas of disconnect between progressive policies and the reality for some people with disabilities in Brazil. Research evidences barriers to health services, including affordability, particularly related to transport, physical inaccessibility of health facilities, and attitudinal and communication barriers^10,23^ which, in turn, limit autonomy. The COVID-19 response also exposed system and service level gaps; we found no evidence that disabled people were represented in COVID-19 coordination groups (or current pandemic preparedness structures). This may have contributed to findings by Sakellariou et al (2020), that while recommendations on disability-inclusive response were published in Brazil, these didn’t translate into formal government policy and there was an over-emphasis on individual level responsibility rather than addressing social structures.^29^ Some government communication on COVID-19 included sign-language interpretation. However, this was likely insufficient; concerns were raised about difficulties faced by deaf people accessing needed information. People with disabilities fared worse in many aspects during the pandemic, including in Brazil,^21,30–32^ it is essential to learn from this and ensure that disabled people are represented in development and implementation of future disaster preparedness plans.

Disability-disaggregated health data are critical for identifying inequalities, stimulating action and monitoring impact. Our analysis highlighted data gaps as well as opportunities in Brazil. The National Health Surveys include internationally recognised questions for generating disability statistics, enabling disability disaggregation of data on health, although we were unable to find evidence of these analysis to date. Another important opportunity is to include disability indicators within SUS Health Information Systems (or enable linkages with other datasets that include disability data); these cover much of the population and are therefore powerful data sources for identifying and monitoring disability-related inequalities. As an example in the UK Learning Disability Registers, GP practises list their patients with learning disabilities and include this information in their electronic health records. These data have been used to compare healthcare for people with and without learning disability and have been important for advocacy and stimulating action at national level.^33,34^ The identified data gaps on population level rehabilitation and AT needs in Brazil, echo a global situation and increasing attention is being paid to methods for collecting these data.^35,36^ Self-reported need, alone, may be unreliable and population surveys that combine self-report and clinical assessments may generate more accurate data to inform service planning.^37,38^ These are costly, but may be feasible to conduct every 5-10 years to provide robust baseline data and monitor progress.

There are also important opportunities for strengthening health-care worker training on disability in Brazil to address some of the service delivery gaps.^39^ Research in different settings highlights this training should involve participatory methods and peer learning, interacting with and learning from people with disabilities.^39^ The Missing Billions Good practise compendium highlights examples of initiatives implemented in other settings, that could be drawn upon, including a Disability-inclusive Nursing Practise Handbook in Germany and rights based disability inclusive health training in Tanzania.^40^

The Missing Billion Health Systems framework was found to be feasible, relevant and comprehensive. The framework was straightforward to use, indicators were generally clear and feasible and most information was in the public domain. We identified some areas for improvement including inclusion of indicators on affordability and on barriers that influence autonomy (e.g. information and attitudinal barriers) and the need to improve scoring metrics or guidance for some of the indicators. In general, the indicators have a greater focus on presence/quantity rather than quality which deserves some attention in terms of interpreting scores. For example, for indicator 7.1 (training of medical doctors on disability); while technically there is training content on disability in medical and non-medical modules (required for a score of 0.66), the actual content is limited and superficial, therefore the score could be misleading.

### Strengths and limitations

The Missing Billions framework provided a structured approach for a comprehensive assessment of disability inclusion in the health system in Brazil. In terms of limitations, we did not engage with Ministry of Health or OPD representatives in completing the indicators. However, their voices were represented in our wider research, which informed the assessment, as well as the formulation of recommendations. It is essential that future health system assessments are led by the Ministry of Health, as is the intention for this framework. Detailed guidelines for this process are available online.^11^. We focused on the public sector for this assessment, however a substantial proportion of the Brazilian population use private healthcare. We did not conduct a comprehensive or systematic literature review as it was not feasible to do this for all indicators. It is therefore possible that some information is missing, for example on disability disaggregated health outcomes.

## Conclusion

The assessment of disability inclusion in the health system, in Brazil, highlighted promotive factors operating at system level in terms of governance, financing and leadership. It also highlighted need and opportunities for strengthening data and evidence, healthcare worker training on disability and accessibility and availability of health services. These actions must be centred on and informed by people with disabilities and underpinned by intersectoral policies to ensure factors influencing health access, outside of the health system (e.g. transport, urban planning), are also addressed.

## Data Availability

All data produced in the present study are available upon reasonable request to the authors

