## Supplemental Tables for "Disability inclusion in the Brazilian health system: results of a health system assessment"

**Supplementary Table 1: Health systems indicators, measure and scoring indicators**

| Indicator and definition | Information required | Scoring methodology |
|---|---|---|
| **1. GOVERNANCE** | | |
| 1.1 Ratification of UNCRPD | Yes / No<br>Evidence of it being actioned, e.g., dedicated budget, action plans and initiatives | Ratified and evidence of action e.g., dedicated budget (1)<br>Ratified with no evidence of action (0.5)<br>No (0) |
| 1.2 Existence of national law protecting rights of persons with disabilities to health | Yes/No<br>National law includes:<br>1) Law prohibits discrimination in healthcare<br>2) Law requires reasonable accommodation for people with disabilities | National law exists without meeting any of the 2 requirements (0.33)<br>With each requirement 0.33 is added to the score |
| 1.3 Existence of national policy or decree on health for persons with disabilities | Yes/No<br>Policy ensures<br>1) General healthcare services for persons with disabilities<br>2) Access to Rehabilitation, other specialists and assistive technology services<br>3) Policy includes measures to implement these services | National policy exists without meeting any of the 3 requirements (0.25)<br><br>With each requirement met 0.25 is added to the score |
| 1.4. Inclusion of people with disabilities in National Health Sector Plan(s) | Yes/No<br>Plan includes:<br>1) Actions and targets for general health care for persons with disabilities (not only prevention of disability)<br>2) Actions and targets for specialist health services for persons with disabilities<br>3) Basic statistics about persons with disabilities and health<br>4) Monitoring and evaluation indicators on disability as part of overall framework for the health sector | People with disabilities are included in National Health Sector Plan(s) without meeting any of the 4 requirements (0.2)<br><br>With each requirement met 0.2 is added to the score |
| 1.5 Inclusion of people with disabilities in National disease plan (e.g., HIV, rare diseases, hepititus) | Yes/no and description<br>Inclusion of people with disabilities in National disease plan (e.g., HIV, rare diseases, hepatitis) | Yes (1)<br>No (0) |

| | | |
|---|---|---|
| 1.6 Cross-ministry taskforce or structure to coordinate work on disability inclusion | Yes/No and which ministry is driving it. Cross ministry governance includes: 1) Department of Health | Yes, for taskforce existing (0.5), additional 0.5 score if Ministry of Health included |
| **2. LEADERSHIP** | | |
| 2.1 MoH Leadership:<br>Existence of a focal point/team in MoH that's responsible for ensuring health access for people with disabilities | Yes/No with description of responsibility for disability inclusion, and title of role/team | There is a role/team responsibility for disability inclusion (1)<br>No (0) |
| 2.2 National health sector coordination: Formal representation of persons with disabilities in highest-level health sector coordination structures | Yes, and title of structure/group<br>No | Yes (1)<br>No (0) |
| 2.3 Pandemic preparedness structure: Formal representation of people with disabilities (individuals or OPD) in national taskforce (e.g. COVID | Yes/No | Yes (1)<br>No (0) |
| **3. HEALTH FINANCING** | | |
| 3.1 Disability Inclusion Budget: Budget (MoH or devolved level) for role/department in MoH working on disability inclusion | Yes/No, description includes if the budget is at the federal/decentralised | Yes, at the federal or decentralized level (1) No (0) |
| 3.2 Reimbursement adjustment for services provided to patients with disabilities | Yes/no<br>For example, there is a national health insurance reimbursement or there is adjusted capitation rates for people with disabilities | Yes, there is a national health insurance and reimbursement for some people with disabilities (1)<br>No adjustments (with any financing mechanisms) (0) |
| 3.2 Funding for AT/rehabilitation in MoH (or devolved levels) budget | Yes/No, % of annual MoH budget | Yes (1)<br>No (0) |

| 4. **DATA AND EVIDENCE** | | |
|---|---|---|
| 4.1 Maturity of disability and health data collection | Yes/No<br>How was is disability and health data gathered?<br>- National census/survey;<br>- Healthcare register of people with disabilities;<br>- Health information records tag people with disabilities (electronic integrated system) | Data is collected through health information records tagging people with disabilities (1)<br>There is a national register for people with disabilities connected to health data (0.67)<br>National survey/census asks disability questions (0.33)<br>Disability and health data is not collected (0) |
| 4.2 Quality of disability and health data collection method | 1) Data collection method is valid<br>2) Data collection is recent - in the last 10 years<br>3) Data is nationally representative 4) 5+ impairment types are covered | Each criteria scores 0.25 points |
| 4.3 Maturity of disability and health data usage | 1) Disability health data that is collected is analysed and published 2) Findings from the data are used to inform program and policy change | 1 - Data is analysed and reported and used to direct policy and program change<br>0.5 - Data is analysed and published<br>0 - Data is not analysed and reported |
| 4.4 Quality of disability and health data usage method | | Each criteria scores 0.25 points |

**Supplementary Table 2: Health service delivery - indicators, measure and scoring indicators**

| Indicator and definition | Information required | Scoring methodology |
|---|---|---|
| **5. AUTONOMY AND AWARENESS** | | |
| 5.1: OPDs advocate on the right to health for persons with disabilities with government and NGO delivery partners | Yes/No - | Yes(1)<br>No (0) |
| 5.2 People with disabilities report autonomy and awareness about health access | Yes/No<br>If the following exist:<br>Quantitative survey (in < 10 years) persons with disabilities were asked about autonomy and awareness about health (in comparison to people without disabilities) OR<br><br>Qualitative data published (in<10 years) in a peer-reviewed journal on reported autonomy and awareness on health | Yes(1)<br>No (0) |
| 5.3 Health information is available in accessible formats | The number of accessibility formats available for the main national health information website e.g., easy read text, web page read out, sign interpretation of video/tv messages, braille, information for care givers | Yes: 2 or more accessibility formats (1)<br>No: less than 2 accessibility formats (0) |
| **6. AFFORDABILITY** | | |
| 6.1 Health coverage – People with disabilities are fully covered for free healthcare through social health insurance, tax-based system, provision as part of disability allowance or any other stipulations | All healthcare is covered / Healthcare is partially covered / No | Yes - There is disability allowance that is available to people with moderate to severe disabilities (1)<br>Yes - There is a disability allowance available for some people living in the country (0.5)<br>No (0) |
| 6.2 Transport subsidy available disabled people and public | Yes/No<br>Yes/No Hospital/health center dedicated public transport services | Yes - there is subsidised transport and facility dedicated services (1)<br>Yes - there is subsidised transport |

| | | |
|---|---|---|
| transport can help travel to medical care | | but not facility dedicated services (0.5)<br>No (0) |
| 6.3 Disability allowance available to cover healthcare fees not covered by existing insurance or tax-based systems, e.g. travel to clinics, assistive technologies | Yes/No<br>Groups and/or regions that have the allowance available | Yes (1)<br>Partial coverage (0.5)<br>No (0) |
| 6.4 Any co-pays for services in either health insurance or taxation based systems are waved for persons with disabilities | Yes/No | Yes (1)<br>No (0) |
| **7. HUMAN RESOURCES** | | |
| 7.1 Training of medical doctors: Information about disability delivered as part of the national curricula for medical schools/colleges | Yes/No<br>Requirements:<br>1) Training content covers medical and non-medical modules<br>2) The training is part of the core curriculum (not voluntary) | Yes without meeting any of the requirements (0.33)<br>With each requirement met 0.33 is added to the score |
| 7.2 Training of nurses: Information about disability delivered as part of the national curricula for nurses/nursing colleges | Yes/No<br>Requirements:<br>1) Training content covers medical and non-medical modules<br>2) The training is part of the core curriculum (not voluntary | Yes without meeting any of the requirements (0.33)<br>With each requirement met 0.33 is added to the score |
| 7.3 Training of CHW: Information about disability delivered as part of the national CHW training curricula | Yes/No<br>Requirements:<br>1) Training content covers medical and non-medical modules<br>2) The training is part of the core curriculum (not voluntary | Yes without meeting any of the requirements (0.33)<br>With each requirement met 0.33 is added to the score |
| 7.4 People with disabilities are represented in the health workforce | % of medical doctors that have disability | Yes - representation is in line with or greater than disability prevalence of the working age population - (if not known for the country assume 2% for LMIC, 4% |

| | | |
|---|---|---|
| | | HIC) (1)<br>No (0) |
| 7.5 Satisfaction: People with disabilities report that they feel well treated by health workers | If the following exist<br>1) In a quantitative survey from within the last 10 years persons with disabilities were asked about satisfaction with health worker services (in comparison to people without disabilities) OR<br>2) Qualitative data published in the last 10 years in a peer-reviewed journal on reported satisfaction | Yes - in a quantitative survey from within the last 10 years persons with disabilities were asked about satisfaction with health worker services (in comparison to people witouth disabilities) (1)<br>OR<br>Yes - qualitative data published in the last 10 years in a peer-reviewed journal on reported satisfaction (1)<br>Both (1)<br>No (0) |
| **8. HEALTH FACILITIES** | | |
| 8.1 Existence of national accessibility standards | Yes/No | Yes (1)<br>No (0) |
| 8.2 Accessibility audit of health facilities has been undertaken in the last 10 years with requirements | Accessibility audit of health facilities has been undertaken in the last 10 years with requirements:<br>1) Results are published<br>2) It is mandatory for all facilities to meet the accessibility standards, consequences when it is not reached | Yes - (0.33)<br>Additional requirements 0.33 each<br>No (0) |
| **9. ASSISTIVE TECHNOLOGY AND REHABILITATION** | | |
| 9.1 National assessment on AT or rehabilitation (e.g. STAR or RATA) done in the last 10 years | Yes/No<br>If yes, provide: 1) description of National Assessment of Assistive technology 2) National representativeness of assessment 3) Date of assessment 4) Key findings from the last assessment | Yes (1)<br>No (0) |
| 9.2 Coordination mechanism cross-Ministry for rehabilitation services | Yes<br>No<br>N/A - only 1 ministry responsible for AT/rehabilitation | Yes (1)<br>No (0)<br>N/A - only 1 ministry responsible |

| | | |
|---|---|---|
| and AT where more than 1 ministries involved | | |
| 9.3 Trained workforce available to provide rehabilitation services and | # of physiotherapists/ 1,000,000 population Includes occupational therapist, audiologist, speech and language, optometrist, Rehabilitation physician, clinical psychologist | 1 - LMIC: Above 30/1,000,000 population<br>HIC: Above 300/1,000,000<br>0 - Below the threshold for population |

**Supplementary Health outputs and outcomes - indicators, definition and status**

| Indicator and definition | Metric |
|---|---|
| **10. EFFECTIVE SERVICE COVERAGE** | |
| 10.1 Modern contraception coverage: Women who demand is satisfied with modern method of contraception disaggregated by disability | % of women with disabilities, compared to % of overall women |
| 10.2 ART coverage: People with HIV receiving ART, disaggregated by disability | % of people with disabilities that have coverage, compared to coverage of people without disabilities |
| 10.3: DPT coverage: Children aged 12-23 months who have received diphtheria-tetanus-pertussis vaccine (DTP3), disaggregated by disability | % of children with disabilities, compared to % of overall children |
| 10.4 Refractive error coverage: People with refractive error have coverage of glasses | % of those with need who have glasses (e.g. from RAAB survey) |
| 10.5 NCD coverage: People with diabetes on treatment OR people with hypertension on treatment, disaggregated by disability | % of people with disabilities, compared to people without disabilities |
| **11. HEALTH STATUS** | |
| 11.1: Overall mortality rate, disaggregated by disability | Deaths per 100 000 population; people with disabilities compared to people without disabilities |
| 11.2 Prevalence of diabetes OR hypertension among persons aged 18+ years, disaggregated by disability | People with disabilities, compared to people without disabilities |
| 11.4: Prevalence of HIV, disaggregated by disability | % of people living with HIV among adults aged 15–49; people with disabilities compared to people without disabilities |
| 11.5 Prevalence of overweight and obesity among persons aged 18+ years, disaggregated by disability | % of all population with disabilities, compared to population without |
| 11.6: Wasting: prevalence of children wasted) disaggregated by disability | |

**Table 4. Key Laws/Policies on access to healthcare for people with disabilities**

| DOCUMENT | MAIN CONTENT | SUMMARY |
|---|---|---|
| **Law/Policy** | **Specification on access to healthcare for people with disabilities** | **Key points related to disability and health** |
| National Policy on Comprehensive Health Care for Persons with Disability (2023)<br>ORDINANCE NO. 1,526, Ministério da Saúde.<br>https://www.in.gov.br/en/web/dou/-/portaria-gm/ms-n-1.526-de-11-de-outubro-de-2023-516446366.<br>Accessed 30 Nov 2023. | Rehabilitation of the Disabled Person, the protection of their health and the prevention of diseases that determine the appearance of disabilities, through the development of articulated actions among the various sectors and the effective participation of society. | • Ordinance to develop National Health Policy for Persons with Disabilities that will:<br>○ Aim at the rehabilitation of people with disabilities, to encourage inclusion<br>○ Protect the health of people with disabilities<br>○ Prevent diseases that result in disability<br>○ Promote participation, collaboration and action of government and non-governmental organisations<br>• Ministry of Health must adjust their plans, programmes and<br>• activities in line with the new policy developed |
| National Plan for the Rights of Persons with Disabilities (November 2023): Decree Nº 11.793,<br>https://www.planalto.gov.br/ccivil_03/_ato2023-2026/2023/decreto/d11793.htm<br>Accessed 30 Nov 2023 | Promote the access to education, health care, social inclusion, and the accessibility for the disabled person. | • Decree to enact the National Plan for the Rights of Persons with Disabilities in line with the CRPD<br>• The plan aims to:<br>○ Prevent causes of disability<br>○ Expand the health care network for people with disabilities<br>○ Improve the standard and quality of health care facilities, including rehabilitation facilities<br>○ Promote access, development and innovation in assistive technology<br>• The bodies involved in implementation must make available their policies, programmes and actions, with information on their budget allocation, as well as the result of their activities<br>• Municipality, State and Federal level agree to collaborate to achieve the targets, prioritizing the inclusion and exercise of rights of people with disabilities |

| | | |
|---|---|---|
| | | • Local bodies may be established to monitor execution of the plan<br>• Government bodies may sign agreements, cooperation plans, etc. with public or private entities, in order to achieve the targets<br>• The plan will be funded by budget allocation from the Federal Government, resources from the implementing bodies and other sources available from agreement with public and private entities |
| Care Network for People with Disabilities within the SUS/ Ministerial Ordinance 793 and Ministerial Ordinance 835<br><br>Brazil. 2012. Portaria Nº 835. Brasília (DF): Ministério da Saúde; Gabinete do Ministro. https://bvsms.saude.gov.br/bvs/saudelegis/gm/2012/prt0835_25_04_2012.html, accessed 05 august 2021.<br>Brazil. 2012. Portaria Nº 793. Brasília (DF): Ministério da Saúde; Gabinete do Ministro. https://bvsms.saude.gov.br/bvs/saudelegis/gm/2012/prt0793_24_04_2012.html, accessed 05 august 2021 | The Care Network for the Disabled Person is organized in: I - Primary care; II- Specialized Attention in Hearing, Physical, Intellectual and Visual Rehabilitation, Ostomy and in Multiple Deficiencies; III- Hospital care and emergency service.<br>Institutes financial incentives for the construction, renovation or expansion of physical location and orthopedic workshop service, as well as for the acquisition of equipment and other permanent materials. | **Ordinance No 835**<br>• Ministry of Health established financial incentives for investment in Specialised Care Component of the Care Network for People with Disabilities, within the scope of the Unified Health System<br>• Finances given for the renovation and expansion of the headquarters of the Specialised Care in Rehabilitation Component, including the orthopedic workshop<br>• Physical facilities must comply with accessibility standards<br>• Rehabilitation centers must:<br> o Have a single record for each patient, with complete history of condition<br> o Have a qualified multidisciplinary team<br> ▪ Physician<br> ▪ Physiotherapist<br> ▪ Speech therapist<br> ▪ Occupational therapist<br> ▪ Social worker<br> ▪ Nurse<br> o Provide 40 hours per week of dental services exclusively for people with disabilities<br> o Have an administrative team |

| | | |
|---|---|---|
| | | ○ If there is visual rehab, hire a rehabilitation expert and technician in mobility<br>• The budget for the resources in this plan shall be borne by the Ministry of Health<br>• The Ministry of health will form a working group to review financing of hearing services and assistive products<br><br>**Ordinance No 793**<br><br>• Establishes the Care Network for people with disabilities within the scope of Unified Health System, across primary care, specialized care in disability rehabilitation and tertiary/emergency care<br>• The general objectives of the Care Network:<br>○ Expand access and improve care for people with disabilities<br>○ Promote link of people with disabilities to points of care<br>○ Guarantee integration and articulation of health networks in a region<br>• The specific objectives of the Care Network:<br>○ Promote healthcare for people with disabilities<br>○ Develop actions for prevention and early intervention<br>○ Expand assistive product provision (specifically orthoses, prostheses)<br>○ Promote rehabilitation and reintegration of people with disabilities (access to work, housing, etc.)<br>○ Promote training for health professionals<br>○ Develop intersectoral action for health promotion and prevention with government and civil society<br>○ Produce and provide information on people with disabilities rights, prevention, care measures, and services available |

| | | |
|---|---|---|
| | | ◦ Organise the demanded of the Care Network<br>◦ Develop monitoring and evaluation of quality services<br>• Ministry of Health will provide support and coordination of the implementation and evaluation, through Municipality Health Departments<br>• Primary care will have:<br>◦ Family Health Support Centres<br>◦ Dental care<br>◦ Promotion and early identification in perinatal care<br>◦ Follow-up with high risk newborns<br>◦ Health education focused on prevention<br>◦ Publish Basic Care guidance for health professionals<br>◦ Develop community projects for inclusion and improved quality of life for people with disabilities<br>◦ Monitoring and health care of people with disabilities at home<br>◦ Support families of people with disabilities<br>◦ Support and provide guidance on health for the education sector<br>• Specialized care will:<br>◦ Have specialised rehab services, specialised dental services, orthopedic workshop<br>◦ Provide comprehensive care<br>◦ Provide accessible information<br>◦ Promote relationship between people with disabilities and health team<br>◦ Adapt services for people with disabilities<br>◦ There are numerous clauses as to the operation of specialized care, including collaboration with primary care, promoting social inclusion, working with families, supporting education sector |
| Brazilian Inclusion Law/ Law 13,146/2015 | Comprehensive health care for people with disabilities is ensured at all levels of complexity, | • The Law is based on the CRPD |

| | | |
|---|---|---|
| Brazil. 2015. Lei Nº 13.146. Brasília (DF): Presidência da República; Secretaria-Geral; Subchefia para Assuntos Jurídicos. http://www.planalto.gov.br/ccivil_03/_ato2015-2018/2015/lei/l13146.htm, accessed 05 august 2021. | through the Unified Health System, guaranteeing universal and equal access. | • Assessment of disability will be carried out by a multi-disciplinary team, accounting for impairment, activity limitation, participation restriction, and personal/environmental factors<br>• Holds general policy statements on equality/discrimination, including right to reproductive rights<br>• PWD will receive priority care in all institutions and services, with access to information provided, resources available, etc.<br>• Right to rehabilitation. There should be:<br>o Early diagnosis and intervention<br>o Measures to compensate for functional limitation<br>o Offer of network of services, at different levels of complexity<br>o Services close to the home of people with disabilities, including rural areas, respecting the Health Care Networks<br>o Rehabilitation services will guarantee: Accessibility, Assistive technology, Training for professionals<br>• Right to health:<br>o Comprehensive healthcare guaranteed on equal basis to all<br>o Participation of people with disabilities in developing and implementing health policies<br>o Care to follow ethical standards, including dignity and autonomy<br>o Professionals supporting people with disabilities must receive continuous training<br>o Public and private health services must ensure:<br>▪ Early diagnosis and intervention<br>▪ Rehabilitation<br>▪ Home care and outpatient treatment<br>▪ Vaccination campaigns |

| | | |
|---|---|---|
| | | ▪ Psychological care (including for family and carers)<br>▪ Respect for gender and sexual orientation of people with disabilities<br>▪ Sexual and reproductive care, including assisted fertilization<br>▪ Accessible information<br>▪ Prevention services<br>▪ Training strategies for teams<br>▪ Guidance for carers<br>▪ Assistive product provision<br>○ Prevention strategies should include:<br>▪ Monitoring in pregnancy and postpartum<br>▪ Nutrition support<br>▪ Immunization<br>▪ Neonatal screening<br>▪ Identification and control of high-risk pregnancy<br>○ Private insurance must guarantee people with disabilities the same services as all customers<br>○ When care cannot be provided at home, people with disabilities must be given transport and accommodation for services elsewhere<br>○ people with disabilities hospitalized are allowed a carer or companion and their stay must be supported by institution<br>○ Discrimination in health is prohibited<br>○ Facilities must be accessible<br>○ Violence against people with disabilities will be addressed by policy and Public Ministry, as well as Council on Rights of Persons with Disabilities<br>• Vocational training |

| | | |
|---|---|---|
| | | ○ Professional qualification must occur in conjunction with public and private networks, especially health<br>• Various statements related to right to transport, right to sport/leisure, right to social assistance, right to work, etc. that can be connected to health (although health is not specifically mentioned in the statements)<br>• Accessibility provisions detailed, including details for building codes, universal design, access to information, etc.<br>• PWD guaranteed access to assistive technology<br>○ Strategies so support access, procedures, research, and resources to be renewed every 4 years<br>• Public power must promote scientific research and innovation designed to improve quality of life for people with disabilities<br>• Provisions for supported decision-making provided<br>○ Process by which a person with a disability elects at least 2 (two) reputable people, with whom they maintain ties and who enjoy their trust, to provide support in decision-making on acts of civil life |
| PNAB/ Ministerial Ordinance 2,436<br><br>Brazil. 2017. Portaria Nº 2,436. Brasília (DF): Ministério da Saúde; Gabinete do Ministro. https://bvsms.saude.gov.br/bvs/saudelegis/gm/2017/prt2436_22_09_2017.html, accessed 05 august 2021. | Ensure adequate infrastructure and good conditions for the operation of Basic Health Units, guaranteeing space, furniture, and equipment, as well as accessibility for people with disabilities, in accordance with current regulations. | • National Primary Care Policy<br>• Primary care is the main gateway and communication centre of the Health Care Network<br>• Basic care free of charge to all people<br>○ Exclusion based on disability is prohibited<br>• Principles in primary care:<br>○ Universality<br>○ Equity<br>○ Person-centred care<br>○ Coordination of care<br>○ Community participation<br>○ Plus others (no mention of disability in any)<br>• Government to ensure accessible infrastructure<br>• Lots of general clauses, but few specific to disability |
